# Anatomic Variations and Contemporary Operative Management of Popliteal Artery Aneurysms

**DOI:** 10.64898/2026.03.10.26348057

**Authors:** Tiffany R. Bellomo, Guillaume Goudot, Natalie Sumetsky, Soumith Sanka, Srihari K. Lella, Brandon Gaston, Shiv Patel, Nikolaos Zacharias, Anahita Dua

**Author notes:** **Corresponding Author:** Tiffany R Bellomo MD, Division of Vascular and Endovascular Surgery Massachusetts General Hospital, 15 Parkman St, WACC-440, Boston, MA 02114. Oral presentation at the 20th annual academic surgical congress 2025 (VSI: ASC 2025).

## Abstract

**Introduction:** Popliteal artery aneurysms (PAAs) are the most common peripheral arterial aneurysm and carry substantial risks of limb loss. Both open and endovascular repair are widely used, yet optimal patient selection remains uncertain. We evaluated institutional operative practices and examined associations between aneurysm morphology, procedural approach, and major adverse limb events (MALE).

**Methods:** We conducted a retrospective cohort study at a tertiary care center to identify patients with PAAs from 2008–2022. Chart review confirmed aneurysm presence and captured demographics, comorbidities, medications, aneurysm characteristics, and operative details. Cox proportional hazards models were used to evaluate time to MALE defined as reintervention or amputation.

**Results:** Among 330 PAAs, median follow-up was 7.4 months (IQR 3.4–12.7). Open repair comprised 79% (250/330), most often a medial approach (75%, 187/250) with autologous vein conduit (65%, 162/250). Open-repair patients were younger than endovascular (69 vs 74 years; p=0.006) with similar cardiovascular profiles. Indications differed by approach, with aneurysm size >20 mm most common for open repair (35.2%, 87/250) and mural thrombus most common for endovascular repair (33.3%, 24/80). MALE occurred in 30.3% (100/330). In univariate analyses, clopidogrel use was associated with increased MALE risk (HR 1.74, 95% CI 1.17–2.59; p=0.006), while descending aortic aneurysm was associated with decreased risk (HR 0.47, 95% CI 0.23–0.92; p=0.029). Operative approach, aneurysm diameter, and thrombus burden were not associated with MALE, and findings were unchanged after multivariable adjustment.

**Conclusions:** MALE risk was comparable across operative strategies and aneurysm morphologies, suggesting that aneurysm size and thrombus burden alone should not preclude consideration of either open or endovascular repair.

## INTRODUCTION

Popliteal artery aneurysms (PAAs) are the most common form of peripheral arterial aneurysms and carry a significant risk of limb-threatening complications^1^. These complications will develop in 31% to 68% of patients with PAA over a lifetime, including thrombosis, distal embolization, acute or chronic limb ischemia, and amputation^2^. Therefore, timely repair of these PAAs is recommended once the aneurysm size or patient symptoms reach the thresholds outlined in the clinical practice guidelines from the Society for Vascular Surgery (SVS)^3^.

Although both open and endovascular approaches have been used to repair PAAs, open repair via a medial approach with autologous great saphenous vein bypass has traditionally shown superior long-term patency, lower occlusion rates, and fewer reinterventions in meta-analyses^1^. However, open repair may be associated with greater perioperative morbidity, wound complications, and longer hospital stays^4^. In contrast, endovascular repair offers shorter operative duration, lower short-term complication rates, and decreased length of hospital stay^5^. Endovascular repair of PAAs is increasingly used in older patients or those with significant comorbidities^5^. However, endovascular repair with a stent could result in kinking, fracture, and in-stent thrombosis^2^.

The optimal management strategy for PAAs varies institutionally and remains a subject of debate, guided by individualized patient considerations. There is uncertainty about the influence of not only procedural approach, but also anatomic factors, such as aneurysm diameter and intraluminal thrombus burden, on postoperative outcomes and risk of major adverse limb event (MALE)^6^. Therefore, the aim of this study was to describe institutional procedural approaches and investigate anatomic characteristics that may be associated with MALE.

## METHODS

### Study Population

The Mass General Brigham Human Research Committee Institutional Review Board approved the study protocol, and patient consent to participate was waived (IRB # 2019P003163). Data were derived from the Massachusetts General Brigham (MGB) Research Patient Data Registry (RPDR), a multi-institutional repository of clinical information, as previously described^7^. Briefly, the RPDR database was queried for all patients with a diagnosis code for “Aneurysm of the artery of lower extremity” (ICD9 442.3/ ICD10 I72.4) from the inception of the database in 2008 to 2022. Manual chart review of each patient was performed by authors T.R.B. and B.G. to confirm the presence of a PAA, defined as greater than 1cm on duplex ultrasound (DUS), consistent with the standard definition of an aneurysm^3^. If there was a discrepancy, senior author A.D. made a final determination. Patients with connective tissue disorders, including Ehlers-Danlos syndrome, Marfan syndrome, and Loeys-Dietz syndromes, were excluded. All patients who underwent an intervention for repair were included in the analysis (Supplemental Figure 1).

### Demographics

Baseline demographics and comorbidities were collected pre-operatively, including age, race, anticoagulation status, smoking history, cardiovascular comorbidities, and other aneurysms. Age was defined at the time of diagnosis prior to the operative intervention. Sex was obtained from fixed categories of female and male. Racial background was self-identified from fixed categories and collapse into White vs non-white. Current smoking was defined smoking at the time of pre-operative assessment. Hypertension was defined as systolic blood pressure ≥130 mmHg, diastolic blood pressure ≥80 mmHg, or prescription of an antihypertensive medication. Hyperlipidemia was defined as total cholesterol ≥200 mg/dL or statin prescription. Diagnoses of cardiovascular comorbidities, including coronary artery disease (CAD), diabetes, dialysis, atrial fibrillation, and deep venous thrombosis immediately prior to the operative intervention. Antiplatelet prescription of aspirin and clopidogrel, or anticoagulation prescription of direct oral anticoagulation (DOAC), warfarin, or enoxaparin, were also ascertained immediately prior to the operative intervention. Concurrent aneurysms were identified through review of operative reports and clinic notes, with categorization by vascular territory (abdominal, ascending, descending aorta; carotid; iliac; other).

### Ultrasound Characteristics

DUS characteristics of PAAs were abstracted by a Registered Physician in Vascular Interpretation certified physician for noninvasive vascular interpretation (G.G.). who was blinded to ultrasound reports. The abstracted characteristics were obtained as previously described^8^. The definitions of each abstracted characteristic were as follows: surface transversal area: the surface of the popliteal aneurysm in cross-section, with inclusion of the arterial wall, from one DUS still image. Internal surface area was defined as the surface of the popliteal aneurysm in cross section, without inclusion of the arterial wall, from one DUS still image. Patent channel area was defined as the surface area of the circulating arterial lumen, free of thrombus, calculated from a single DUS image. Thrombus area was defined as the area within the artery occupied by thrombus material, calculated by subtracting the area of the true lumen from the area of the entire aneurysm. Anteroposterior diameter was defined as the largest diameter in any still image measured in the anterior to posterior dimension. Transverse diameter was defined as the largest diameter in any still image measured in the transverse dimension. Largest diameter was defined as the maximum diameter annotated in either anteroposterior or transverse dimensions. Percent thrombus was calculated by dividing the thrombus area by the area of the aneurysm by multiplied by 100.

### Operative Characteristics

The primary exposure was operative intervention, classified as endovascular or open surgery. Endovascular intervention was defined as procedures in which the primary component of the initial operation was performed endovascularly. Open intervention was defined as occurrence of an open surgical bypass or interposition graft. Amputation was not considered a primary intervention. The primary indication for intervention was defined according to the reason for surgery listed in the operative report or postoperative note. Emergent intervention was defined as cases documented as emergent operations in the operative report. Return to the operating room (OR) was defined as reoperation within 72 hours to address a procedure-related complication and was not considered a reintervention. The primary outcome was MALE, defined as any repeat open or endovascular procedure performed to restore or maintain patency of a previously constructed bypass graft or stent, including procedures for graft stenosis, occlusion, or recurrent ischemia, and major amputation. Both the indication and type of MALE were recorded.

### Statistical Analysis

Patient demographics were stratified by procedural approach of endovascular or open intervention, with each PAA treated as a separate event. Measurements from DUS were expressed as median and interquartile range (IQR) due to the nonparametric distribution of data. Univariate Cox proportional hazards regression models were used to quantify the relationship between clinical pre-operative characteristics and MALE. Multivariable Cox proportional hazards models were constructed using variables significant in univariate analysis, along with age, race, sex, and initial intervention, which were included a priori based on clinical relevance. The proportional hazards assumption was assessed using Schoenfeld residuals and was not violated (Global p=0.914). Statistical analyses were performed using STATA, version 15.1 (StataCorp, College Station, Texas). In all analyses, statistical significance was determined using an α = 0.05 threshold (p<0.05).

## RESULTS

### Demographic and Anatomic Characteristics

A total of 330 PAAs underwent operative intervention, where 168 were bilateral aneurysms in 84 patients and 162 were unilateral aneurysm in 162 patients for a total of 246 patients. Follow up time was a median of 7.4 months (IQR 3.4, 12.7) after the procedural approach. Patients who underwent open intervention were younger (median of 69 vs 74, p=0.006) than those who underwent endovascular intervention. Otherwise, cardiovascular comorbidities and medications were similar between both groups (Table 1). Most PAAs on first presentation were greater than 2cm (87.3%, 288/330) and contained mural thrombus (61.5%, 203/330). Patients with a PAA on first presentation had symptoms of claudication (52.4%, 173/330), followed by acute limb ischemia (ALI) (21.5%, 71/330), chronic limb ischemia (CLI) (8.2%, 27/330), and least commonly rupture (1.8%, 6/330).

**Table 1.** Characteristics and demographics of 246 patients with 330 popliteal artery aneurysms that underwent repair stratified by procedure approach. P-values reflect comparisons between open and endovascular repair and were calculated using Fisher’s exact test or chi^2 test, as appropriate, for categorical variables, and the Kruskal-Wallis test for continuous variables. CLI, chronic limb-threatening ischemia; ALI, acute limb ischemia.

| Demographics<br>n (%) | Open<br>(n = 250) | Endovascular<br>(n = 80) | Total<br>(n = 330) | P-value |
| --- | --- | --- | --- | --- |
| Age (median, IQR) | 69 (61–77) | 74 (65–81) | 70 (62–78) | 0.006* |
| Race (White) | 235 (94.0%) | 74 (92.5%) | 309 (93.6%) | 0.632 |
| Sex (Male) | 242 (96.8%) | 78 (97.5%) | 320 (97.0%) | 1.000 |
| Laterality |  |  |  | 0.190 |
| Left | 121 (48.4%) | 32 (40.0%) | 153 (46.4%) |  |
| Right | 129 (51.6%) | 48 (60.0%) | 177 (53.6%) |  |
| Ever smoker | 195 (78.3%) | 57 (71.2%) | 252 (76.6%) | 0.194 |
| Current smoker | 25 (10.0%) | 10 (12.5%) | 35 (10.6%) | 0.535 |
| Hypertension | 224 (89.6%) | 71 (88.8%) | 295 (89.4%) | 0.83 |
| Hyperlipidemia | 210 (84.0%) | 66 (82.5%) | 276 (83.6%) | 0.752 |
| Coronary artery disease | 126 (50.4%) | 46 (57.5%) | 172 (52.1%) | 0.269 |
| Diabetes | 79 (31.6%) | 24 (30.0%) | 103 (31.2%) | 0.788 |
| Dialysis | 20 (8.0%) | 4 (5.0%) | 24 (7.3%) | 0.464 |
| Atrial fibrillation | 80 (32.0%) | 27 (33.8%) | 107 (32.4%) | 0.771 |
| Deep venous thrombosis | 86 (34.4%) | 22 (27.5%) | 108 (32.7%) | 0.252 |
| Antiplatelet |  |  |  |  |
| Aspirin | 230 (92.0%) | 72 (90.0%) | 302 (91.5%) | 0.576 |
| Clopidogrel | 115 (46.0%) | 43 (53.8%) | 158 (47.9%) | 0.227 |
| Anticoagulation |  |  |  |  |
| DOAC | 48 (19.2%) | 19 (23.8%) | 67 (20.3%) | 0.379 |
| Warfarin | 130 (52.0%) | 45 (56.2%) | 175 (53.0%) | 0.507 |
| Enoxaparin | 97 (38.8%) | 28 (35.0%) | 125 (37.9%) | 0.542 |
| Concurrent Aneurysms |  |  |  |  |
| Abdominal aorta | 167 (66.8%) | 43 (53.8%) | 210 (63.6%) | 0.035 |
| Descending aorta | 42 (16.9%) | 11 (13.8%) | 53 (16.2%) | 0.501 |
| Ascending aorta | 22 (8.9%) | 5 (6.2%) | 27 (8.2%) | 0.458 |
| Carotid artery | 5 (2.0%) | 1 (1.2%) | 6 (1.8%) | 1.000 |
| Iliac artery | 92 (36.8%) | 29 (36.2%) | 121 (36.7%) | 0.929 |
| Other | 82 (32.8%) | 25 (31.2%) | 107 (32.4%) | 0.797 |
| Symptoms on Presentation |  |  |  |  |
| Size > 2cm | 222 (88.8%) | 66 (82.5%) | 288 (87.3%) | 0.141 |
| Mural thrombus | 153 (61.2%) | 50 (62.5%) | 203 (61.5%) | 0.835 |
| Occlusion | 63 (25.2%) | 21 (26.2%) | 84 (25.5%) | 0.851 |
| Claudication | 134 (53.6%) | 39 (48.8%) | 173 (52.4%) | 0.45 |
| CLI | 18 (7.2%) | 9 (11.2%) | 27 (8.2%) | 0.25 |
| ALI | 52 (20.8%) | 19 (23.8%) | 71 (21.5%) | 0.576 |
| Rupture | 4 (1.6%) | 2 (2.5%) | 6 (1.8%) | 0.635 |

Among popliteal artery aneurysms with available preoperative DUS measurements, no significant differences in anatomic characteristics were observed between those treated with open compared to endovascular repair (Supplemental Table 1). The median aneurysm diameters were 23.5 mm (IQR 19.6–33.6) in the AP dimension, 25.2 mm (20.4–36.2) in the transverse dimension. The areas measured included a total surface area of and 490 mm² (327-1,016), an internal surface area of 462 mm² (283-990), a patent channel area of 151 mm² (75.4–248), and a thrombus area of 276 mm² (113.5–712). Percent thrombus was high in both groups, measuring 69% (43–89) in PAAs treated with open repair and 75% (50–83) in those treated with endovascular intervention.

### Operative Factors

The majority of PAAs (79%, 250/330) underwent open intervention through a medial incision (75%, 187/250) with an autologous vein conduit (65%, 162/250) (Table 2). Of the PAAs that underwent endovascular intervention, all repairs were performed via percutaneous access, and 8 (13.8%, 8/80) underwent thrombolysis. Open interventions were booked less frequently as urgent procedures (19.8% vs 32.4%, p=0.025). The most common indication for intervention, was size greater than 20mm in open interventions at 35.2% (87/250) compared to mural thrombus presence in endovascular interventions at 33.3% (24/80) (Table 2). Eleven popliteal artery aneurysms required an acute return to the operating room, most commonly for acute thrombosis (5 aneurysms) following open surgical repair; two aneurysms in the endovascular group also required reoperation for compartment syndrome.

**Table 2.**
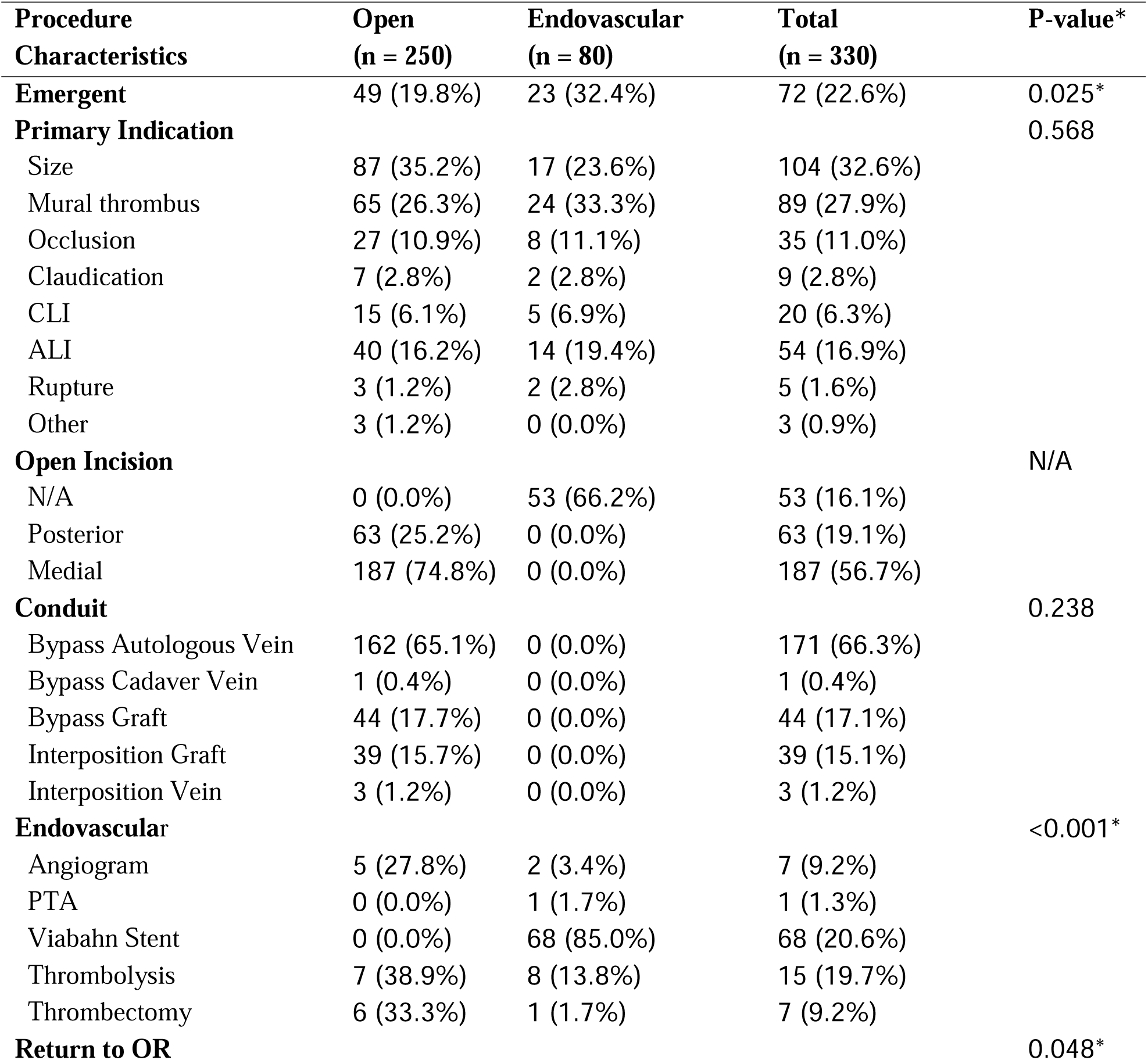
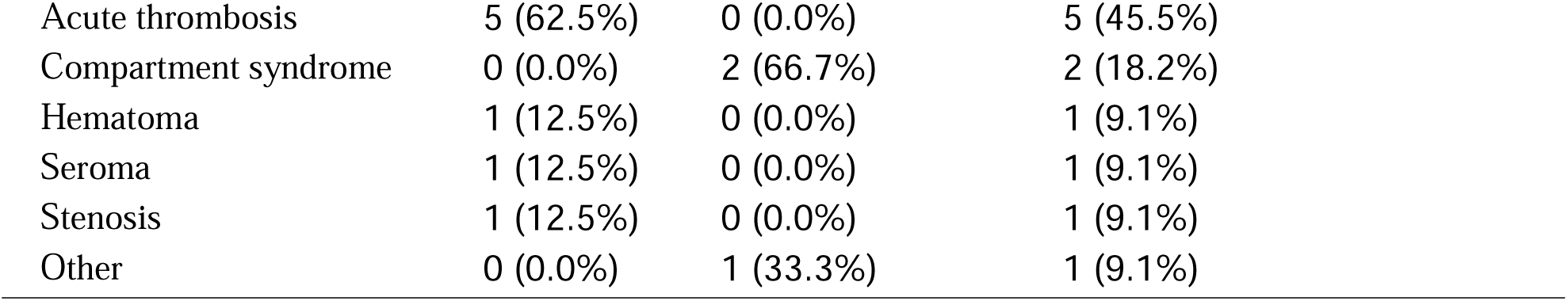
Procedural characteristics of 330 popliteal artery aneurysms among 246 patients undergoing repair, analyzed per aneurysm and stratified by operative approach (open vs endovascular). Data are presented as n (%) using aneurysm-level denominators (open, n = 250; endovascular, n = 80; total, n = 330). P-values reflect comparisons between open and endovascular repair and were calculated using Fisher’s exact test or the chi^2 test for categorical variables. Sections not applicable to a given approach are indicated as N/A. Abbreviations: CLI, chronic limb-threatening ischemia; ALI, acute limb ischemia; PTA, percutaneous transluminal angioplasty.

### Outcome Measures

A total of 100 PAAs had a MALE event, including 77 that underwent initial open intervention and 23 that underwent initial endovascular intervention. The median time to MALE was longer after open repair (15 months; IQR 5–80) compared with endovascular repair (6 months; IQR 1–34) (Table 3). The most common indication for reintervention was occlusion in both procedural approaches, with 25.0% (19/77) in open and 52.2% (12/23) in endovascular initial approaches. Reinterventions after open approach included bypass with vein (22.1%, 17/77), bypass with graft (19.5%, 15/77), stenting (14.3%, 11/77), percutaneous angioplasty (13.0%, 10/77), thrombectomy (13.0%, 10/77), other combinations of interventions (13.0%, 10/77), thrombolysis (3.9%, 3/77), and drug-coated balloon angioplasty (1.3%, 1/77). Reinterventions after endovascular approach included bypass with vein (30.4%, 7/23), bypass with graft (17.4%, 4/23), thrombectomy (17.4%, 4/23), stenting (17.4%, 4/23), percutaneous angioplasty (8.7%, 2/23), other combinations of interventions (4.3%, 1/23), and thrombolysis (4.3%, 1/23). Ultimately, 24 PAAs that underwent intervention resulted in amputation. The median time to amputation was similar between open (n=15, median 6 months; IQR 1–78) and endovascular interventions (n=9, median 2 months; IQR 1–87). Amputation level did not differ significantly between groups (p = 0.920), with above-knee amputation being the most common (46%, 11/24), followed by toe (29%, 7/24), below-knee (13%, 3/24), and transmetatarsal amputations (13%, 3/24).

**Table 3.**
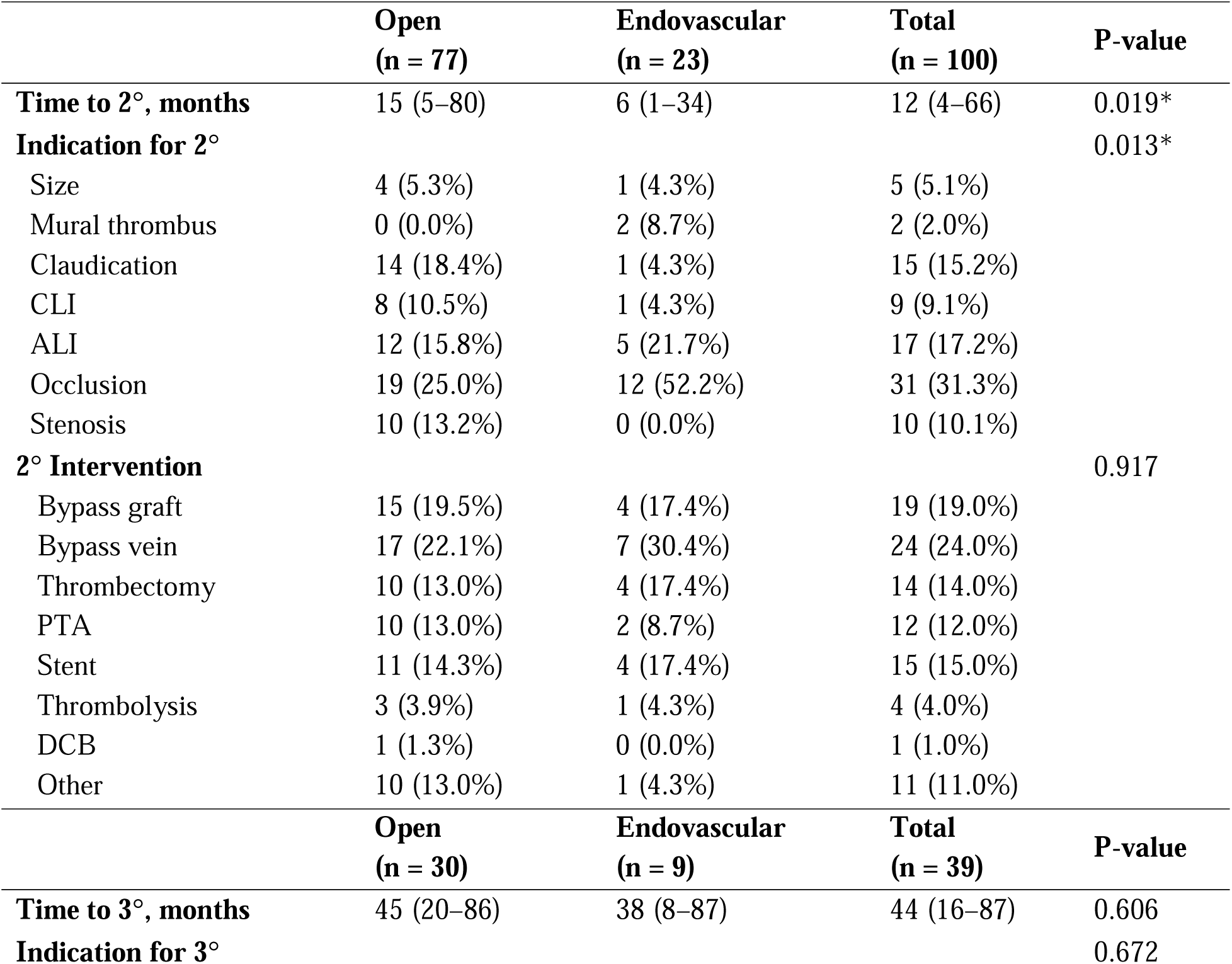
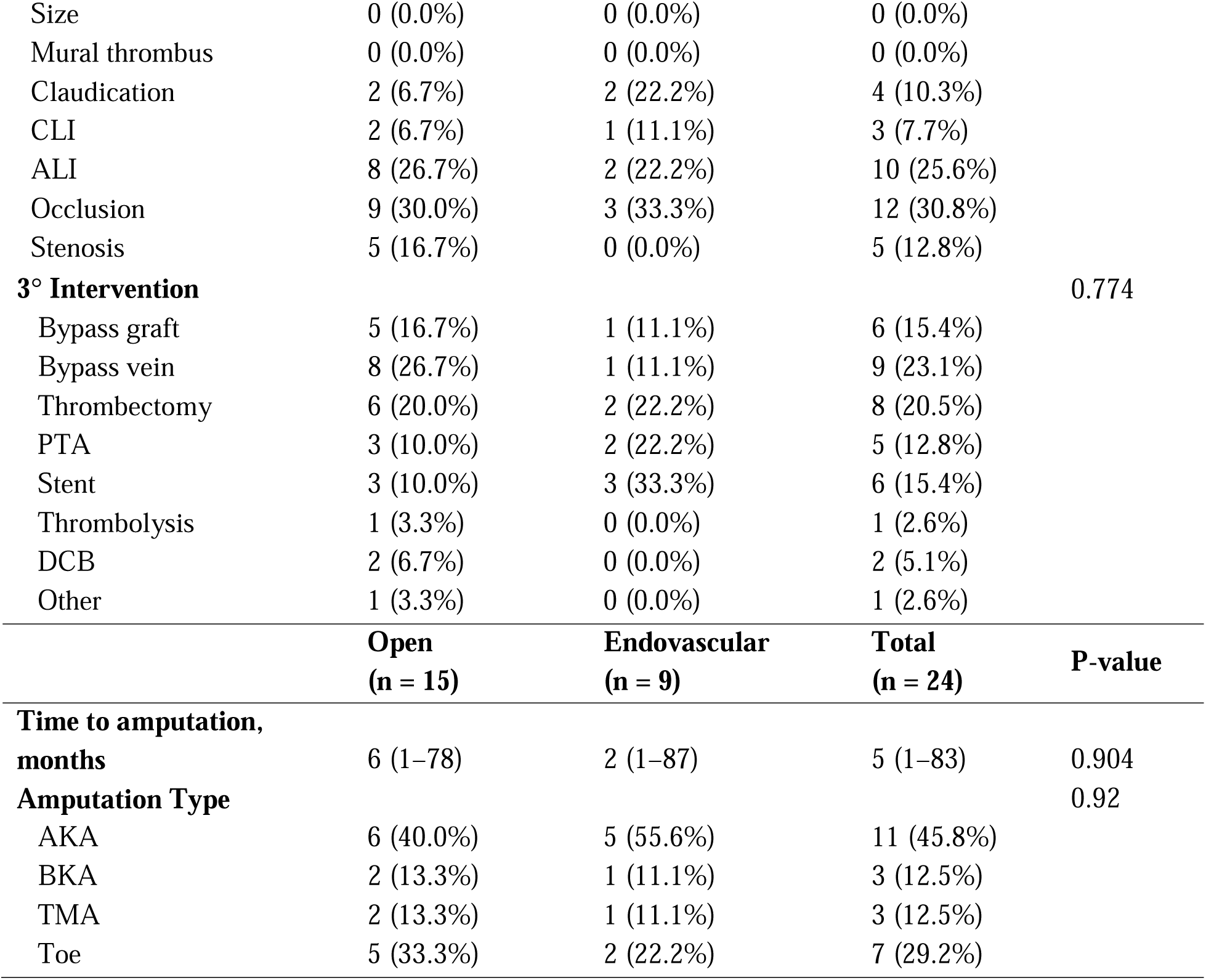
Major adverse limb events outcomes following repair of popliteal artery aneurysms, analyzed per aneurysm and stratified by operative approach. Secondary (2°) and tertiary (3°) reinterventions are reported for aneurysms experiencing each event, with denominators reflecting the number of aneurysms at risk within each subgroup. Continuous variables are reported as median (interquartile range). P-values compare open versus endovascular repair and were calculated using the Kruskal-Wallis test for continuous variables and Fisher’s exact test or the chi squared test for categorical variables. Abbreviations: 2°, secondary; 3°, tertiary; CLI, chronic limb-threatening ischemia; ALI, acute limb ischemia; PTA, percutaneous transluminal angioplasty; DCB, drug-coated balloon; AKA, above-knee amputation; BKA, below-knee amputation; TMA, transmetatarsal amputation.

In univariate time-to-MALE analyses of demographic characteristics and comorbidities, clopidogrel use was associated with an increased risk of MALE (HR 1.74, 95% CI 1.17–2.59; p=0.006), whereas aspirin and anticoagulation use were not significantly associated with MALE (Supplemental Table 2). The presence of a concurrent descending aortic aneurysm was associated with a reduced risk of MALE (HR 0.47, 95% CI 0.23–0.92; p=0.029). Other concurrent aneurysms were not significantly associated with MALE, although point estimates suggested hazards less than 1. In addition, open surgical repair, compared with endovascular intervention, was not significantly associated with MALE (HR 0.70, 95% CI 0.44–1.10, P = 0.124). In multivariable analysis adjusting for clopidogrel use, presence of a descending aortic aneurysm, and prespecified covariates (age, race, and sex), open repair compared to endovascular repair was not significantly associated with MALE (HR 0.61, 95% CI 0.38–1.01; p=0.054) (Figure 1; Supplemental Table 3). When analyses were restricted to popliteal artery aneurysms with available preoperative duplex ultrasound measurements, neither percent thrombus (HR 3.24, 95% CI 0.58–18.04; p=0.179) nor largest aneurysm diameter (HR 1.00, 95% CI 0.97–1.02; p=0.708) was associated with time to MALE in unadjusted analyses (Supplemental Table 4) or after adjustment for the same covariates (Supplemental Table 5).

## DISCUSSION

Although both open and endovascular repair have acceptable outcomes, the optimal surgical strategy for PAAs varies institutionally and there is uncertainty about the influence of anatomic factors on MALE^6^. This descriptive study utilized a large cohort of 330 PAAs that underwent operative intervention at a tertiary care center with manually annotated DUS images of various anatomic characteristics. The majority of PAAs (79%, 250/330) underwent open intervention through a medial incision (75%, 187/250) with an autologous vein conduit (65%, 162/250). In univariate time-to-MALE analysis, neither percent thrombus nor aneurysm diameter was associated with MALE. The only variable associated with increased risk of MALE was clopidogrel use, which may represent confounding by indication for the prescription. In multivariate analysis, an initial open approach was still not significantly associated with MALE. These findings address a national knowledge gap regarding practice variability in PAA repair and suggest that large aneurysm size or high thrombus burden should not discourage pursuing either open or endovascular repair.

Although SVS guidelines define high risk PAAs as having mural thrombus, poor run off, and a diameter greater than 20 mm^3^, there remains no clear clinical consensus on specific anatomic thresholds for repair. Observational studies consistently report intervention at sizes well above 20 mm, with median diameters of 25 mm in Spain^9^ and 30–32 mm in Sweden and the US^6,10^. Our findings align with this pattern, where PAAs that underwent intervention had a median repair diameter of 25.2 mm. Mural thrombus is also considered a high-risk feature^3^, and prior work has shown it to be a significant predictor of aneurysm sac expansion^11^. This is comparable to our cohort, where thrombus burden was substantial in both groups, with percent thrombus burden at 69% (IQR 43–89) in open repairs and 75% (IQR 50–83) in endovascular repairs.

Both open and endovascular approaches are widely used for PAA repair. Open repair of PAAs can be performed through either medial or posterior incisions^3^. The posterior approach offers excellent exposure for complete sac decompression, whereas the medial approach allows the patient to remain supine and facilitates simultaneous access to inflow and outflow vessels^12^. In our cohort, open repairs were predominantly performed through a medial incision (74.8%), with the posterior approach used in 25.2%. This distribution aligns with national and international trends, including data from the Vascunet registry of 14 countries, where 77.7% of open repairs used a medial approach and 22.3% used a posterior approach^13^. Similar to these studies, most open bypasses in our cohort used autologous vein. In contrast, endovascular repair involves placement of a covered stent to exclude the aneurysm, with variability in practice regarding the need for femoral cutdown. Unlike the POPART study, which reported that most endovascular repairs were performed via femoral cutdown^2^, all endovascular interventions in our study were performed via percutaneous access. Endovascular repair has been increasingly utilized as the definitive repair of PAAs over the years^14^, and has also increased specifically in cases of acute limb ischemia^15^, consistent with the proportion of endovascular to open interventions in our study.

The operative management of PAAs has evolved considerably with the advent of endovascular techniques, yet the optimal procedural approach for PAAs remains a subject of ongoing debate. Several comparative studies suggested advantages to open repair, including higher primary patency in the POPART registry^2^, lower rates of major amputation^14^, and overall lower major adverse limb events in a Vascular Quality Initiative^1,16^ at one year. Despite these early benefits, other studies report no significant differences between open and endovascular repair in primary and secondary patency at two years ^17,18^, or in amputation free survival in the large multi-center PARADE study^9,19^. Consistent with these reports, our study similarly found no significant differences between open and endovascular repair with respect to amputation rates. In univariate and multivariant analysis, we also observed no significant differences in time to MALE between open and endovascular repair in our cohort. Conversely, several studies highlight potential advantages of endovascular repair, including shorter length of stay^4,15^, decreased perioperative morbidity^15^, and higher primary patency^20^ all at one year. A contemporary meta-analysis in 2021 further reflects this mixed evidence^1^. While open repair had lower vessel occlusion rates and fewer reinterventions within 30 days than endovascular interventions, open repair was also associated with lower primary patency at one year.

Recent studies highlight that the determinants of MALE extend beyond the procedural approach itself, underscoring the importance of patient and anatomic characteristics. Consistent with prior literature^9^, our study found that patients undergoing endovascular intervention were more often older; however, this was not associated with an increased risk of MALE^21^. We also observed mixed findings regarding anticoagulation, likely reflecting confounding by indication, with all anticoagulation and antiplatelet use appearing to increase MALE risk. Notably, high-quality evidence guiding optimal anticoagulation therapy for PAA patients remains limited, and no specific regimen has consistently improved outcomes after lower-extremity revascularization^22^. We observed that the presence of a descending aortic aneurysm was significantly associated with a reduced risk of MALE; however, this finding may reflect increased surveillance among patients with aortic aneurysms, as PAAs may not be followed as closely as recommended^8^. Anatomic characteristics have also been investigated, where traditionally larger aneurysm size or greater thrombus burden were thought to be associated with increased risk of post-operative complications and MALE^3,6^. However, in our analysis, both diameter and percent thrombus were not associated with MALE. This aligns with findings from a German cohort of over 300 PAAs, in which aneurysm size was not independently associated with MALE^21^. Although poor tibial runoff is well established as a predictor of reintervention following both open and endovascular repair^23^, our ability to assess tibial vessel patency was limited because preoperative ultrasound imaging often lacked consistent or meaningful documentation.

### Limitations

This study should be interpreted in the context of its limitations inherent to its retrospective design. The Massachusetts General Brigham consists primarily of tertiary referral hospitals, and patients may have received follow-up care at outside institutions, contributing to the relatively short median follow-up observed. The technical parameters of DUS examinations were not standardized, and variability in operator technique may have influenced the measured diameters and thrombus areas. Given the modest sample size and lack of an a priori power calculation, the possibility of type II error cannot be excluded. Surgeon-specific factors, including individual technical proficiency, were not accounted for and may have affected the likelihood of MALE. Additionally, tibial vessel patency was not consistently evaluated preoperatively, limiting assessment of its contribution to MALE risk. Finally, procedures for patients with bilateral PAAs were analyzed independently, which may have introduced correlated outcomes within individuals.

## CONCLUSIONS

In this cohort, operative approach and aneurysm anatomic features, including percent thrombus and diameter, were not significantly associated with MALE. By demonstrating comparable MALE risk across operative strategies and aneurysm morphologies, these findings address a national knowledge gap underlying practice variability in popliteal artery aneurysm repair. These findings suggest that aneurysm size and thrombus burden alone should not preclude consideration of either open or endovascular repair. Future research should focus on identifying specific anatomic and clinical predictors that optimize outcomes for each approach and on establishing long-term comparative data beyond currently available follow-up periods.

## Data Availability

Data that were used in this study are available on request from the senior author (A.D.). Code to perform analyses in this manuscript are available from the authors upon request (A.D.).

## ACKNOWLEDGEMENTS

We thank the participants at our institution for their data used in this study.

## Abbreviations

PAAs: Popliteal artery aneurysms
CLI: Critical limb ischemia
ALI: Acute limb ischemia
TE: Thromboembolic event
PPV: Positive predictive value
SVS: Society for Vascular Surgery
DUS: Duplex ultrasound
STROBE: Strengthening the Reporting of Observational Studies in Epidemiology
MGB: Massachusetts General Brigham
RPDR: Research Patient Data Registry
AP: Anteroposterior
CTA: Computed tomography angiography

---

**Figure 1.** Multivariable Cox proportional hazards model for major adverse limb events following popliteal artery aneurysm repair. Forest plot displaying hazard ratios (HRs) and 95% confidence intervals from a multivariable Cox proportional hazards model evaluating factors associated with major adverse limb events following popliteal artery aneurysm repair (n = 330). Covariates included age, race, sex, presence of a descending aortic aneurysm, initial operative approach (open repair), and antiplatelet therapy variables identified as significant in univariate analyses. The dashed vertical line indicates the null value (HR = 1.0). HRs greater than 1 indicate increased risk, whereas HRs less than 1 indicate reduced risk of adverse limb events. Model discrimination was modest (Harrell’s C = 0.595).

## REFERENCES

1. Beuschel B, Nayfeh T, Kunbaz A, et al. A systematic review and meta-analysis of treatment and natural history of popliteal artery aneurysms. J Vasc Surg. 2022;75(1S):121S–125S.e14.

2. Jung G, Leinweber ME, Karl T, et al. Real-world data of popliteal artery aneurysm treatment: Analysis of the POPART registry. J Vasc Surg. 2022;75(5):1707–1717.e2.

3. Farber A, Angle N, Avgerinos E, et al. The Society for Vascular Surgery clinical practice guidelines on popliteal artery aneurysms. J Vasc Surg. 2022;75(1S):109S–120S.

4. Leake AE, Avgerinos ED, Chaer RA, Singh MJ, Makaroun MS, Marone LK. Contemporary outcomes of open and endovascular popliteal artery aneurysm repair. J Vasc Surg. 2016;63(1):70–76.

5. Tielliu IFJ, Verhoeven ELG, Zeebregts CJ, Prins TR, Span MM, van den Dungen JJAM. Endovascular treatment of popliteal artery aneurysms: results of a prospective cohort study. J Vasc Surg. 2005;41(4):561–567.

6. Jergovic I, Cheesman MA, Siika A, et al. Natural history, growth rates, and treatment of popliteal artery aneurysms. J Vasc Surg. 2022;75(1):205–212.e3.

7. Bellomo TR, Goudot G, Lella SK, et al. Percent thrombus predicts Popliteal artery aneurysm related limb threatening events. Ann Surg. 2025;282(6):1134–1139.

8. Bellomo TR, Goudot G, Gaston B, et al. Popliteal artery aneurysm ultrasound criteria for reporting characteristics. Vasc Med. 2024;29(1):58–63.

9. Serrano Hernando FJ, Martínez López I, Hernández Mateo MM, et al. Comparison of popliteal artery aneurysm therapies. J Vasc Surg. 2015;61(3):655–661.

10. Kropman RHJ, van Santvoort HC, Teijink J, et al. The medial versus the posterior approach in the repair of popliteal artery aneurysms: a multicenter case-matched study. J Vasc Surg. 2007;46(1):24–30.

11. Ascher E, Markevich N, Schutzer RW, Kallakuri S, Jacob T, Hingorani AP. Small popliteal artery aneurysms: are they clinically significant? J Vasc Surg. 2003;37(4):755–760.

12. Taurino M, Filippi F, Ficarelli R, et al. Different approaches in popliteal artery aneurysm management. Eur Surg. 2013;45(4):221–226.

13. Grip O, Mani K, Altreuther M, et al. Contemporary treatment of popliteal artery aneurysms in 14 countries: A vascunet report. Eur J Vasc Endovasc Surg. 2020;60(5):721–729.

14. Wrede A, Wiberg F, Acosta S. Increasing the elective endovascular to open repair ratio of popliteal artery aneurysm. Vasc Endovascular Surg. 2018;52(2):115–123.

15. Satam K, Aboian E, Cardella J, et al. The management of patients with popliteal artery aneurysms presenting with acute limb ischemia. J Vasc Surg. 2023;78(2):506–513.

16. Eslami MH, Rybin D, Doros G, Farber A. Open repair of asymptomatic popliteal artery aneurysm is associated with better outcomes than endovascular repair. J Vasc Surg. 2015;61(3):663–669.

17. Pulli R, Dorigo W, Castelli P, et al. A multicentric experience with open surgical repair and endovascular exclusion of popliteal artery aneurysms. Eur J Vasc Endovasc Surg. 2013;45(4):357–363.

18. Ronchey S, Pecoraro F, Alberti V, et al. Popliteal artery aneurysm repair in the endovascular era: Fourteen-years single center experience. Medicine (Baltimore). 2015;94(30):e1130.

19. Troisi N, Bertagna G, Saratzis A, et al. Elective surgical repair of popliteal artery aneurysms with posterior approach vs. Endovascular exclusion: Early and long term outcomes of multicentre PARADE study. Eur J Vasc Endovasc Surg. 2025;69(1):110–117.

20. Slysz JT, Tian L, Zhao L, Zhang D, McDermott MM. Effects of supervised exercise therapy on blood pressure and heart rate during exercise, and associations with improved walking performance in peripheral artery disease: Results of a randomized clinical trial. J Vasc Surg. 2021;74(5):1589–1600.e4.

21. Freytag H, Kapalla M, Berg F, et al. Bypass patency and amputation-free survival after popliteal aneurysm exclusion significantly depends on patient age and medical complications: A detailed dual-center analysis of 395 consecutive elective and emergency procedures. J Clin Med. 2024;13(10):2817.

22. Zavgorodnyaya D, Knight TB, Daley MJ, Teixeira PG. Antithrombotic therapy for postinterventional management of peripheral arterial disease. Am J Health Syst Pharm. 2020;77(4):269–276.

23. Zaghloul MS, Andraska EA, Leake A, et al. Poor runoff and distal coverage below the knee are associated with poor long-term outcomes following endovascular popliteal aneurysm repair. J Vasc Surg. 2021;74(1):153–160.

